# Sporadic Oncocytic Tumors with Features Intermediate between Oncocytoma and Chromophobe Renal Cell Carcinoma: Comprehensive Clinico-Pathological and Genomic Profiling

**DOI:** 10.1101/2020.05.28.20115931

**Authors:** Yajuan J Liu, Cigdem Ussakli, Tatjana Antic, Yuhua Liu, Yu Wu, Lawrence True, Maria S Tretiakova

## Abstract

**Background:** Morphology, clinical behavior, and genomic profiles of renal oncocytoma (RO) and its malignant counterpart chromophobe renal cell carcinoma (ChRCC) are distinctly different. However, there is a substantial group of sporadic oncocytic tumors with peculiar hybrid phenotypes as well as a perplexing degree of morphologic and immunohistochemical overlap between classic RO and ChRCC with eosinophilic cytoplasm. The aim of this study is to provide detailed characterization of these hybrid tumors.

**Design:** Thirty eight sporadic oncocytic neoplasms with ambiguous morphology from two institutions were reviewed by 4 pathologists. CKIT positivity was used as a selection criterion. We correlated CK7 and S100A1 immunostaining and detailed morphologic features with cytogenetic profiles. DNA from the FFPE tissues was extracted and analyzed using Cytogenomic Microarray Analysis (CMA) to evaluate copy number alterations and ploidy.

**Results:** CMA categorized cases into 3 groups: RO (N=21), RO variant (N=7) and ChRCC (N=10). Cytogenetic RO had either no CNA (48%) or loss of chromosome 1p, X or Y (52%). RO-variant had additional chromosomal losses [-9q, –14 (n=2), –13] and chromosomal gains [+1q (n=2), +4, +7 (n=2), +13, +19, +20, and +22]. ChRCC were either hypodiploid with numerous monosomies (40%) or hypotetraploid with multiple relative losses (60%). RO, RO-variant and ChRCC groups differed significantly in tumor architecture (p<0.01), stroma (p=0.013), presence of nuclear wrinkling, perinuclear halos and well-defined cell borders in >5% cells (p<0.01), focal cell clearing (p=0.048) and CK7 expression (p<0.02). Pathologic prediction of cytogenetic subtype using only two categories (benign RO or malignant ChRCC) would overcall or undercall up to 40% of tumors that were ChRCC based on cytogenetics. This finding provides the rationale for an intermediate diagnostic category of so-called hybrid tumors (HOCT). HOCT was a heterogeneous group enriched for cytogenetic RO-variant. Other HOCTs have a profile of either RO or ChRCC.

**Conclusions:** Genomic profile allows classification of oncocytic tumors with ambiguous morphology into RO, RO-variant and ChRCC. Several architectural and cytologic features combined with CK7 expression are significantly associated with cytogenetic RO, RO-variant or ChRCC tumors. Doubled hypodiploidy by whole genome endoduplication is a common phenomenon in eosinophilic ChRCC.

Parts of this study were presented in an abstract form at the 104^th^ annual meeting of the United Stated and Canadian Academy of Pathology, Boston, March 21–27, 2015

## INTRODUCTION

Distinguishing oncocytoma (RO) from eosinophilic chromophobe renal cell carcinoma (ChRCC) is often difficult even among expert urologic pathologists, especially in needle biopsies [1–4]. Yet the distinction is important since RO is benign and ChRCC can be not only locally aggressive but can also metastasize. Sporadic hybrid oncocytic/chromophobe tumors (HOCT) represent a poorly understood controversial entity with gross, architectural and cellular features that overlap with both RO and ChRCC that has prominent eosinophilic features [3–6]. Though selected immunostains help address this differential diagnosis, there remain cases incompletely resolved by immunohistochemistry. ChRCC is characterized by multiple chromosomal losses (most frequently chromosomes Y, 1, 2, 6, 10, 13, 17 and 21) [7]. Conversely RO usually has a normal number of chromosomes, occasionally exhibiting genetic abnormalities that include loss of whole chromosome 1 or part of its short or long arm and partial or complete loss of chromosomes 14, 21 and Y [7–11]. We hypothesize that molecular characterization by cytogenomic microarray analysis of problematic oncocytic tumors with a non-iconic morphology and perplexing immunohistochemical profile that overlaps with RO and/or ChRCC can provide a basis for categorizing the majority of these oncocytic tumors.

## MATERIALS AND METHODS

### Case Selection

This retrospective study used cases retrieved from the pathology archives of the University of Washington and the University of Chicago Medical Centers, with approval of Institutional Review Boards. We interrogated the pathology databases of our two institutions, searching for the following categories: “Oncocytic tumor, NOS” or “Oncocytic cell neoplasm” with varying modifiers including “low-grade”, “borderline features”, “unclassified”, “low malignant potential”, “with hybrid features” as well as “Oncocytoma with atypical features” and “Hybrid oncocytic and chromophobe tumor”. Additional selection criteria included CKIT expression by tumor cells and availability of material for further immunohistochemical and molecular studies. We identified 35 cases of sporadic, unifocal, oncocytic neoplasms that met all selection criteria and had an ambiguous morphology, at least focally, preventing making a definitive diagnosis of either classic RO or ChRCC. Three cases of ChRCC with prominent eosinophilia, suggestive of hybrid morphology, were added from the Cancer Genome Atlas (TCGA) cohort [12]. We excluded cases with Birt-Hogg-Dube (BHD) syndrome, succinate-dehydrogenase (SDH) deficiency syndrome, renal oncocytosis, angiomyolipomas, and cases with a history of tuberous sclerosis complex (TSC). None of selected cases had morphologic features and immunohistochemical profiles of such recently described oncocytic tumor entities as eosinophilic, solid and cystic renal cell carcinoma (ESC-RCC), high-grade oncocytic tumor (HOT) or low-grade oncocytic tumor (LOT) [13, 14].

Cases were reviewed by 4 pathologists [MT, AT, CU, LT] to document architectural patterns (nested/organoid, tubulocystic, solid/confluent), quality of stroma (scant, edematous, fibromyxoid, calcified or with osseous metaplasia), presence of hemorrhage, necrosis, fat or vascular invasion, frequent binucleation/multinucleation and cellular pleomorphism. Among atypical features worrisome for malignancy we recorded cell clearing when present in >5% of tumor cells, raisinoid nuclei), perinuclear halos, “vegetable-like” cell membranes, as well as >1 mitoses [2, 6, 7, 15]. For comparison purposes we recorded frequency of various morphologic parameters including tumor architecture, stromal component, cytologic appearance and atypical features which were previously shown useful in distinguishing RO, HOCT and ChRCC [2, 4, 6, 16].

### Immunohistochemistry

The unstained sections were deparaffinized by two xylene rinses followed by two rinses with 100% ethanol. The immunostaining was performed in a CLIA certified diagnostic immunohistochemistry laboratory according to standardized protocol. In brief, antigen retrieval was performed on an automated immunostainer (BondTM, Leica, Germany) using H2 buffer (pH 8.0) for 20 minutes. After rinsing and endogenous peroxidase blocking, the slides were incubated for 25 minutes with a mouse antibodies against CKIT (CD117, polyclonal, 1:250, DAKO/Agilent, cat A4502), CK7 (clone OV-TL, 1:200, DAKO/Agilent cat M7018) and S100A1 (clone 56C6, 1:25, Novocastra, cat NCL-CD10–270). Slides were then rinsed multiple times and incubated for 30 minutes with anti-mouse polymer detection reagent (Refine kit, Leica, Germany). This was followed by multiple rinses, incubation with diaminobenzidine chromogen, and hematoxylin counterstain. Negative controls replaced the primary antibody with non-immune mouse serum. The immunostaining was interpreted as previously described in the study addressing optimization of immunohistochemical profiles in the differential diagnosis of RO from ChRCC [17]. The neoplasm was considered positive when by >50% tumor cells stained diffusely and negative when only single cells and small cell clusters stained.

### Cytogenomic Microarray Analysis (CMA)

All 35 specimens used in this study were archived formalin-fixed paraffin embedded (FFPE) tissue. The methods for DNA isolation from FFPE specimens, genomic microarray analysis using Agilent SurePrint G3 Cancer CGH+SNP 4×180K Array http://www.chem-agilent.com/pdf/5990-9183en_lo_CGH+SNP_Cancer.pdf), and copy number evaluation have been described previously [18].

Six cases with complex copy number aberrations (Figure 2, cases 1, 2, 3, 4, 5, 34) were also analyzed using Illumina Infinium CytoSNP-850K BeadChip to confirm the ploidy status.

Genomic DNA extracted from the FFPE specimen was end-repaired, amplified, and hybridized to the Illumina Infinium CytoSNP-850K BeadChip v1.1 (http://www.illumina.com/content/dam/illuminamarketing/documents/products/datasheets/datasheet_CytoSNP850K_POP.pdf) The microarray was washed, labeled, stained, and scanned with an Illumina iScan as specified by the manufacturer (Illumina Inc, San Diego, CA, USA). Allele and intensity ratio data of the fluorescent signals were generated. Microarray data were analyzed and visualized using Nexus manufacturer (Illumina Inc, San Diego, Copy Number 10.1 (BioDiscovery, Inc. Hawthorne, CA, USA) to identify chromosomal copy number variants and regions of copy number neutral loss of heterozygosity. Genome build GRCh37 was used.

### Statistical analysis

Demographic and clinical characteristics were summarized for each subtype of oncocytic tumor and compared across subtypes using Kruskall-Wallis test for continuous variables (age and tumor size), and chi-square or Fisher exact test for categorical variables (stage, morphologic and immunohistochemical features). Median follow-up time was reported for patients who were alive at the end of follow-up. Results were considered statistically significant only if p≤0.05.

## RESULTS

### Clinicopathologic and immunohistochemical findings

Mean patient age was 59 years with slight predominance of males over females (1.6:1 ratio). All tumors were unifocal with a mean tumor size of 4.8 cm (range 1.8 to 20.6 cm). Twenty-five tumors were treated by partial nephrectomy (65.8%), twelve by radical nephrectomy (31.6%). One patient (2.6%) was not surgically treated. Pathologic stage at surgery distributed as follows: pT1a (65.8%), pT1b (15.8%), pT2a (5.3%), pT2b (5.3%) and pT3a (7.8%). The median follow-up was 56 months (range 2–137). None of the patients had metastases or died of disease. However, tumor recurred in 2 patients, and 3 patients died from other causes.

Morphologically all tumors were composed of round to polygonal tumor cells with densely granular bright eosinophilic cytoplasm, round to oval nuclei with largely inconspicuous nucleoli. Architecturally 32/38 (84%) tumors had a nested/“archipelagenous” or organoid architecture, 21/38 (55%) had tubulocystic areas and 21/38 (55%) had a solid/confluent architecture; the combination of 2 architectural patterns was noted in 20/38 (53%) tumors, whereas all three architectures were present in 8/38 (21%) neoplasms. Edematous paucicellular stroma was present in 19/38 (50%), fibromyxoid in 23/38 (61%), and calcifications and/or osseous metaplasia was observed in 5/38 (13%) tumors. Hemorrhage, prominent, pleomorphic nuclei and multinucleation were seen in 42%, 24% and 32% cases, respectively. Of cytologic features present in >5% of tumor cells we saw 1) nuclear wrinkling in 11/38 (29%), 2) perinuclear halos in 15/38 (40%), 3) well-defined cell borders in 12/38 (32%) and 4) cell clearing in 14/38 (37%) tumors, as well as 5) increased mitotic activity in 3/38 (8%) (Figure 1). Fat invasion was present in 4/38 (11%) cases whereas vascular invasion in 1/38 (3%) case. No tumors in our cohort had areas of necrosis or apoptotic debris.

**Figure 1.**
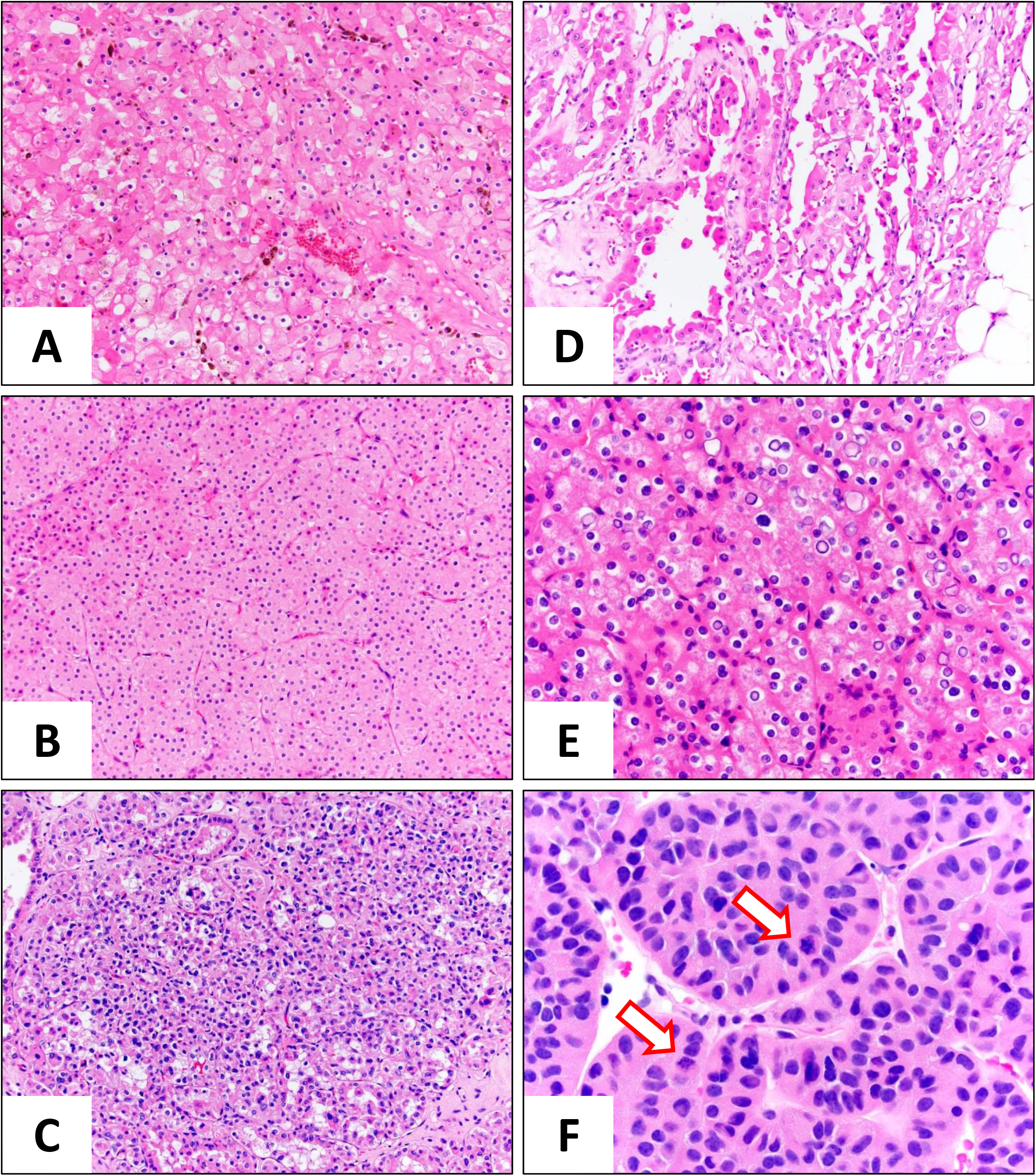
Representative examples of morphologic features of oncocytic renal cell tumors worrisome for malignancy. A – Focal cell clearing and perinuclear halos, case #1 (20x); B – Solid/confluent growth pattern and well-defined cell borders, case #4 (20x); C – nuclear wrinkling (irregular nuclear membranes), pleomorphism and multinucleation, case #29 (20x); D – Tubulocystic architecture and fat invasion in case #12 (20x); E – diffuse perinuclear clearing and frequent binucleation, case #22 (40x); F – Increased mitotic activity (arrows) in case #35 (60x).

CD117 (CKIT) immunoreactivity was invariably positive in the majority of tumor cells. Diffuse positivity of CK7 (>50% cells) was documented in 11/38 (29%) cases. S100A1 cytoplasmic and nuclear expression in the majority of tumors cells was present in 23/32 (72%) cases.

### Prediction of benign versus borderline vs malignant diagnoses

The diagnostic criteria favoring benign RO included one atypical morphologic feature plus an immunoprofile of negative CK7/positive S100A1 (N=20). Eosinophilic variant of ChRCC was favored in cases with >3 atypical features and cases with positive CK7/negative S100A1 immunoprofile (N=7). All other cases with borderline/intermediate features were considered hybrid oncocytic chromophobe tumors (HOCT) (N=11).

### Cytogenetic findings

The criteria of genetic profiles for RO included normal CMA or chromosomal loss –1/1p, –14 (less frequent), –X and/or –Y [7, 10]. RO-variant genetic profiles included a few additional copy number aberrations (mostly gains) in addition to a chromosomal loss –1/1p-, –14, –X and/or –Y in RO. The WHO criteria of genetic profiles for ChRCC was hypodiploidy, including combination of numerous chromosomal losses of one copy of the entire chromosomes –1, –2, –6,–10, –13, –17, –21, and –Y [7].

#### 3.1. Renal oncocytoma (RO) by CMA

Twenty one cases (55%) had genetic profiles characteristic of RO. These included ten cases with normal CMA’s, seven cases with loss of chromosome 1 including an additional loss of the Y chromosome in four cases and of an X chromosome in one case. And there were four cases with loss of portions of the short arm (p) of chromosome 1 (1p-) including one case that also had loss of the Y chromosome (Figure 2).

**Figure 2.**
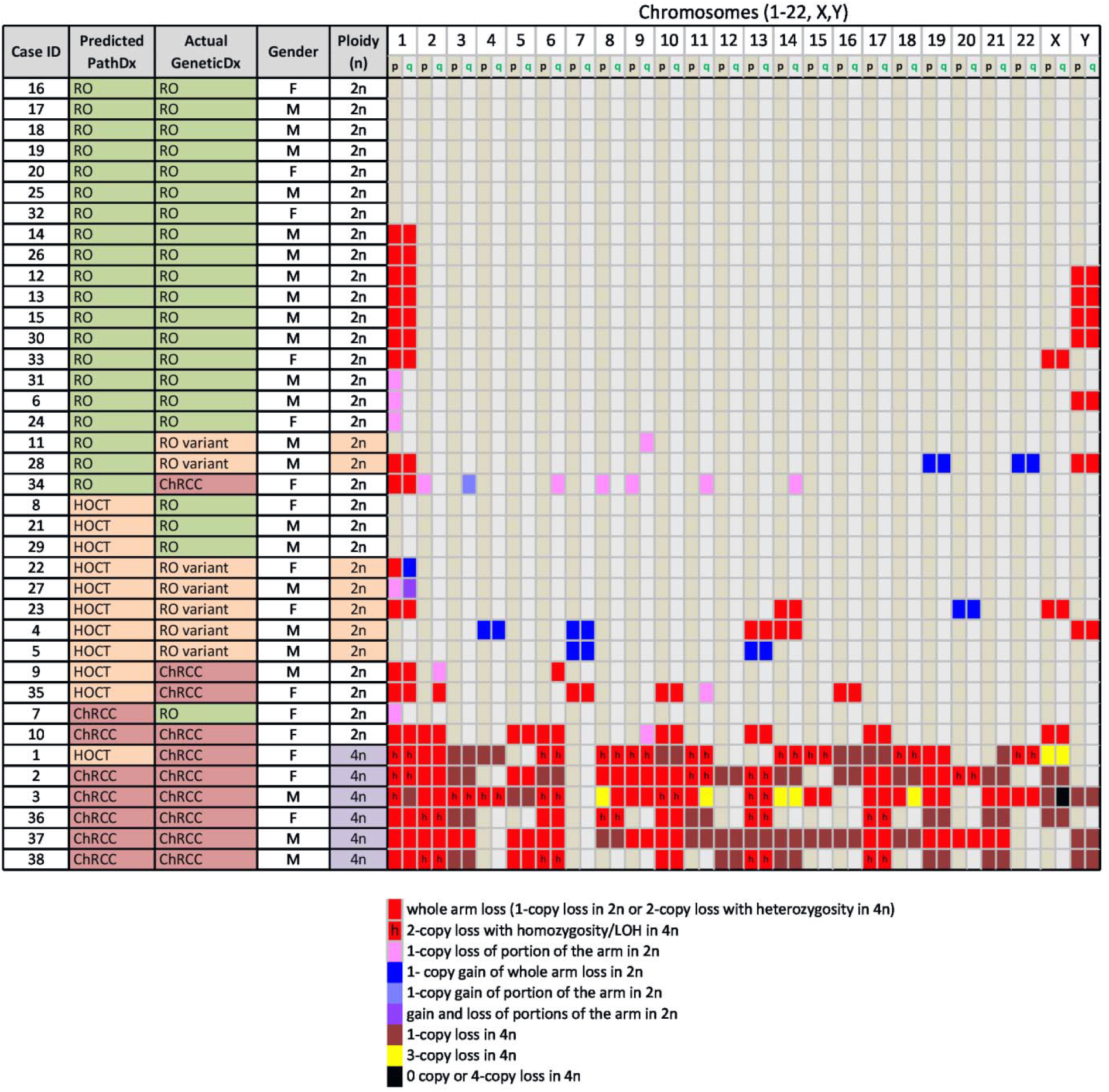
Cytogenetic results (actual) classifying cases as RO, RO-variant or ChRCC compared to favored pathology diagnosis (predicted) RO, HOCT and ChRCC shows match in 28 cases (74%) and mismatch or reclassification in 10 cases (26%).

**Figure 3.**
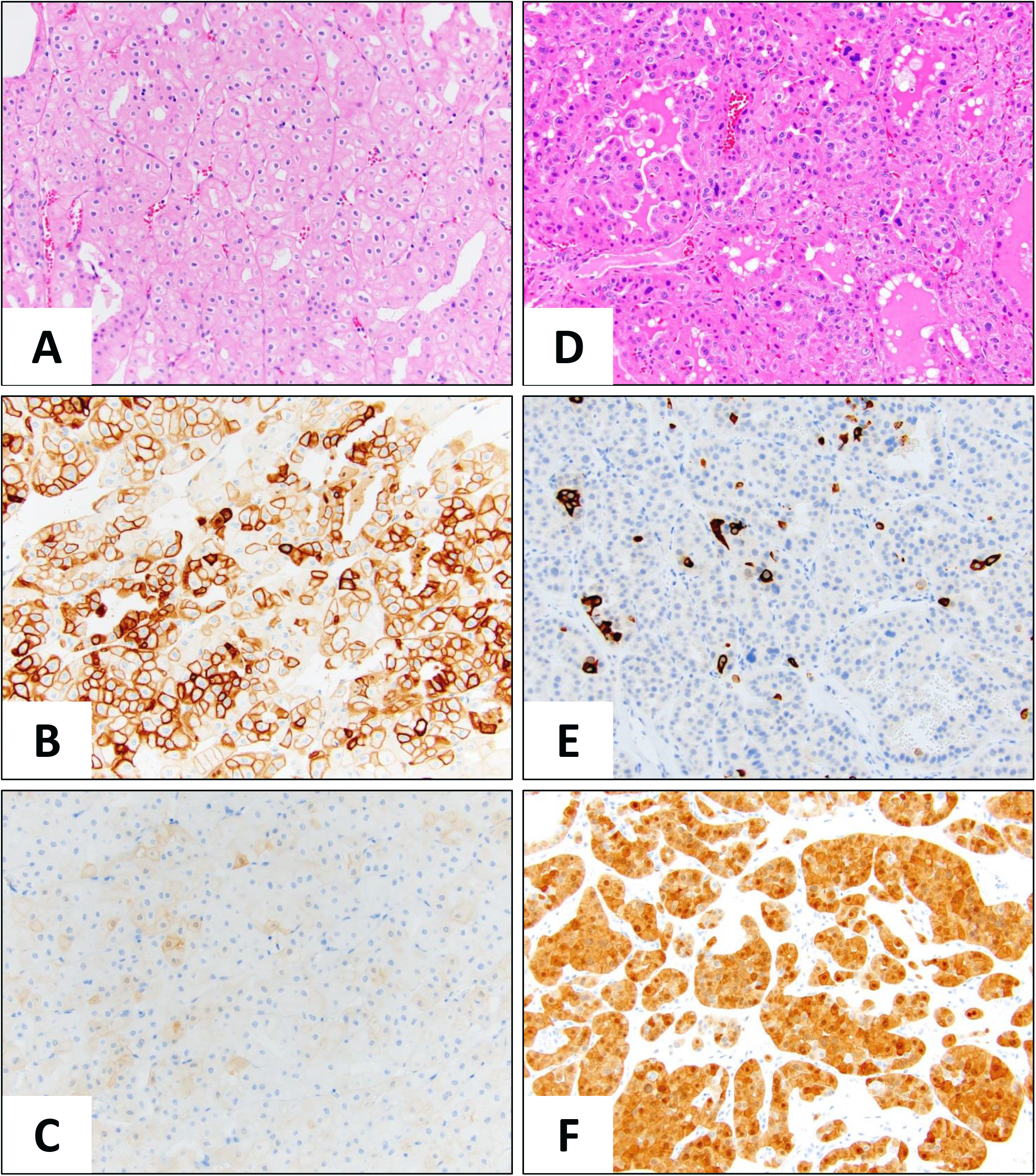
Two cases with major discrepancy between cytogenetic (actual) and pathologic (predicted) diagnoses. A-C: Case #7 with cytogenetic RO profile (loss of 1p), but morphologically (A) most consistent with eosinophilic variant of ChRCC with diffuse CK7 expression (B) and negative for S100A1 (C). D-F: Case #34 with cytogenetic ChRCC profile (loss of 1p, 2p, 6q, 8p, 9p, 11q, 14q), but morphologically (D) most consistent with RO with scattered CK7 positivity (E) and diffuse nuclear and cytoplasmic S100A1 (F); magnification 20x.

#### 3.2. RO-variant by CMA

Seven tumors (18.5%), which had aberrations common in RO’s, such as chromosomal loss –1/1p-, –14, –X, and –Y, also had one to three additional copy number aberrations, which were mostly gains. We interpreted these as RO-variants. These aberrations included gain of 1q in two cases, gain of 7 in two cases; one case each with gain of chromosomes 13, 19, 20 and 22, sole deletion of portion of 9q31.3qter, and gain of chromosome 4 in addition to loss of chromosome 13 (Figure 2).

#### 3.3. Chromophobe RCC by CMA

Ten cases (26.5%) had multiple chromosomal abnormalities including four cases with hypodiploidy, which is characterized as losses of portions or entire chromosomes such as 1, 2, 6, 10, 11, 16, and 17. These changes are genetically consistent with ChRCC. In addition, six cases, including three TCGA cases with eosinophilic ChRCC phenotype, had multiple chromosomal losses due to hypotetraploidy (doubled hypodiploidy). The karyotypes inferred from the CMA results are summarized in Table 1 and Figure 2. The status of chromosomes with 2 copy numbers and loss of heterozygosity (LOH) indicated that one copy of these chromosomes were most likely lost initially in the diploid state (2n) before whole genome endoduplication, while chromosomes with 2 or 3 copy numbers without LOH were due to subsequent loss in the tetraploid state (4n) after whole genome endopduplication event. The most frequently affected chromosomes with two-copy loss in these cases with hypotetraploidy (doubled hypodiploidy) included chromosomes 1 (100%), 2 (100%), 5 (50%), 6 (83%), 8 (67%), 9 (67%), 10 (83%), 11 (50%), 13 (67%), 17 (83%), 19 (67%), X (100% in females), and Y (100% in males). These patterns of chromosomal losses are consistent with ChRCC genetic profiles. The chromosomes with three (one copy loss in 4n) or four copies (no loss in 4n) in these six hypotetraploid ChRCC cases were most frequently chromosomes 3 (67%), 4 (83%), 7 (100%), 12 (100%), 14 (67%), 15 (67%), 16 (100%), 18 (67%), 20 (67%), 21 (67%), and 22 (67%) (Table 1, Supplemental Table 1).

**Table 1.** Karyotypes of six ChRCC with hypotetraploidy (doubled hypodiploidy).

| **Case ID** | **Karyotypes based on CMA results** |
| --- | --- |
| 1 | <4n>61,X,-X,-X,-X,(-1<sup>h</sup>, -2, -6<sup>h</sup>, -8<sup>h</sup>, -9<sup>h</sup>, -11<sup>h</sup>, -14<sup>h</sup>, -15<sup>h</sup>, -18<sup>h</sup>, -19, -22<sup>h</sup>)x2, -3, -4, -10, -16, -17, -21 |
| 2 | <4n>62,XXX,-X,(-1<sup>h</sup>, -2, -5, -8, -9, -10, -11<sup>h</sup>, -13<sup>h</sup>, -17, -19, -20<sup>h</sup>)x2, -3, -6, -12, -14, -16, -18, -21 |
| 3 | <4n>55,del(X)(q13.2)Y,-X,-Y,(-1p<sup>h</sup>, -2<sup>hp</sup>, -3<sup>h</sup>, -4<sup>h</sup>, -6<sup>h</sup>, -8<sup>hp</sup>, -9, -10<sup>h</sup>, -11<sup>hp</sup>, -13<sup>h</sup>, -14<sup>h</sup>, -15, -17, -18<sup>hp</sup>, -19, -21, -22,)x2, -1q, -5, -8p, -11q<sup>p</sup>, -14, -18q<sup>p</sup> |
| 36 | <4n>73,XXX,-X,(-1, -2<sup>h</sup>, -6, -8<sup>h</sup>, -10, -13<sup>h</sup>, -17<sup>h</sup>)x2, -3, -11, -19, -21 |
| 37 | <4n>61,XXY,-Y,(-1, -2, -3, -5, -6, -9, -10, -17, -19, -20, -21)x2, -8, -11, -12, -13, -14, -15, -16, -18 |
| 38 | <4n>72,XXY,-Y,(-1, -2<sup>h</sup>, -5, -6<sup>h</sup>, -10, -13<sup>h</sup>, -17<sup>h</sup>)x2, -3, -12, -14, -19, -21 |

Of note, <sup>h</sup> - homozygosity or loss of heterozygosity; <sup>hp</sup> - homozygosity for portion of the chromosomal arm; <sup>p</sup> – portion of the chromosomal arm (see supplemental table 1 for details).

#### Clinico-pathological features of cytogenetically classified oncocytic tumors

There were no statistically significant differences between the three subtypes (RO, RO-variant and ChRCC) in age, gender, type of surgery, tumor size, stage or outcomes (Table 2). Comparison of architectural patterns showed higher frequency of nested/organoid or so-called “archipelagenous” architecture in RO and RO-variant comparing to ChRCC (p<0.01), whereas solid/confluent architecture was more common in ChRCC and RO-variant, but not in RO (p<0.01). Tubulocystic morphology was more frequent in RO and ChRCC, and less common in RO-variant (p<0.01). Fibromyxoid and edematous stroma was characteristic for the majority of RO and much less common in RO-variant and ChRCC (p=0.014, p=0.013 respectively). Presence of hemorrhage, stromal calcification or osseous metaplasia was not discriminatory between three tumor types. From cytologic features nuclear wrinkling, perinuclear halos and well-defined borders in >5% cells were typical for the majority of ChRCC and common in RO- variant tumors, but not in RO (p<0.01). Focal cell clearing was present in 70% ChRCC, 43% RO-variant tumors and 24% of RO marginally reaching statistical significance (p=0.048). Presence of cellular pleomorphism and bi/multinucleation was not discriminatory between tumor types (p=0.29 and p=0.96, respectively). Among atypical features very few cases showed increased mitotic activity, fat or vascular invasion, and no cases had tumor necrosis/apoptosis, thus did not show statistically significant difference between study groups.

**Table 2.** Demographic and clinico-pathological features of cytogenetically classified oncocytic tumors: classic renal oncocytoma (RO), RO-variant and chromophobe renal cell carcinomas (ChRCC).

| Features | Details | RO | RO-variant | ChRCC | P-value |
| --- | --- | --- | --- | --- | --- |
| **Cases** | N=38 | N=21 | N=7 | N=10 |  |
| **Pt age (mean)** | 59.1 years | 63.2 | 58.3 | 50.8 | 0.09 |
| **M:F ratio** | 1.6:1 | 1.5:1 | 2.5:1 | 0.7:1 | 0.21 |
| **Surgery** | Partial nephrectomy | 18/21 (86%) | 5/7 (71%) | 7/10 (70%) | 0.52 |
|  | Radical nephrectomy | 3/21 (14%) | 2/7 (29%) | 2/10 (20%) |  |
|  | Biopsy | - | - | 1/10 (10%) |  |
| **Tumor size (mean)** | 4.8 cm | 4.3 cm | 3.5 cm | 6.6 cm | 0.21 |
| **Stage** | pT1a | 14/21 (67%) | 6/7 (86%) | 4/9 (44%) | 0.45* |
|  | pT1b | 4/21 (19%) | - | 2/9 (22%) |  |
|  | pT2a | 1/21 (5%) | 1/7 (14%) | - |  |
|  | pT2b | - | - | 2/9 (22%) |  |
|  | pT3a | 2/21 (9%) | - | 1/9 (11%) |  |
| **Follow-up (median)** | 56 months | 59 mo | 24 mo | 67 mo | 0.14 |
| **Outcomes** | No evidence of disease | 13/21 (62%) | 4/7 (57%) | 8/10 (80%) | 0.53 |
|  | Recurrence | 1/21 (5%) | - | 1/10 (10%) |  |
|  | Died of other cause | 2/21 (9%) | 1/7 (14%) | - |  |
|  | Lost to follow-up | 5/21 (24%) | 2/7 (29%) | 1/10 (10%) |  |
| **Architecture** | Nested/archipelagenous | 21/21 (100%) | 6/7 (86%) | 5/10 (50%) | <0.01 |
|  | Tubulocystic | 16/21 (76%) | 1/7 (14%) | 4/10 (40%) | <0.01 |
|  | Solid/diffuse growth | 7/21 (33%) | 5/7 (71%) | 9/10 (90%) | <0.01 |
| **Stroma** | Fibromyxoid | 17/21 (81%) | 3/7 (43%) | 3/10 (30%) | 0.014 |
|  | Edematous | 15/21 (71%) | 2/7 (29%) | 2/10 (20%) | 0.013 |
|  | Calcified/osseous metaplasia | 4/21 (19%) | 1/7 (14%) | - |  |
|  | Hemorrhage | 10/21 (48%) | 2/7 (29%) | 4/10 (40%) | 0.67 |
| **Cytology** | Pleomorphism | 7/21 (33%) | 1/7 (14%) | 1/10 (10%) | 0.29 |
|  | Multinucleation | 7/21 (33%) | 2/7 (29%) | 3/10 (30%) | 0.96 |
|  | Nuclear wrinkling <sup>(A)</sup> | 1/21 (5%) | 4/7 (57%) | 6/10 (60%) | <0.01 |
|  | Perinuclear halos <sup>(A)</sup> | 3/21 (14%) | 4/7 (57%) | 8/10 (80%) | <0.01 |
|  | Well-defined cell borders <sup>(A)</sup> | 1/21 (5%) | 3/7 (43%) | 8/10 (80%) | <0.001 |
|  | Cell clearing <sup>(A)</sup> | 5/21 (24%) | 3/7 (43%) | 7/10 (70%) | 0.048 |
| **“Aggressive” features** | >1 mitoses <sup>(A)</sup> | 1/21 (5%) | - | 2/10 (20%) | 0.18 |
|  | Apoptosis/necrosis <sup>(A)</sup> | - | - | - | - |
|  | Fat invasion | 3/21 (14%) | - | 1/10 (10%) | 0.74 |
|  | Vascular invasion | 1/21 (5%) | - | - | - |
| **Immunostaining** | CK7 (>50% cells) | 3/21 (14%) | 2/7 (29%) | 6/9 (67%) | 0.016 |
|  | S100A1 (>50% cells) | 15/19 (80%) | 4/6 (67%) | 4/7 (57%) | 0.52 |

“-“ – feature not present; \* - stage p-values is comparing stage 1/2 vs 3; <sup>(A)</sup> – Atypical features

Positive expression of CK7 in more than 50% tumor cells was present in 9% of RO, 43% RO-variant cases and 67% ChRCC, yielding statistically significant difference (p=0.016). Diffuse nuclear and cytoplasmic S100A1 positivity was more frequent in RO group and RO-variant groups (80% and 67%) compared to 57% ChRCC cases, but not significantly (p=0.52).

#### Correlation between immunomorphologic and cytogenetic diagnoses

From seven immunomorphologically predicted ChRCC cases (n=7), six were confirmed cytogenetically (86% match), whereas one case had RO cytogenetics (Figure 2, case 34). From twenty predicted RO cases (n=20), seventeen were cytogenetically RO (85% match), whereas two were RO-variants with 9q loss and gain of 19 and 22 (Figure 2, cases 11 and 28) and one tumor had ChRCC cytogenetics (Figure 2, case 7). And finally, in morphologically borderline tumors (HOCT, n=11), three cases (27%) were classified as ChRCC, five as RO-variants (46%) and three as RO (27%) by cytogenetic profiling (Figure 2, Table 3).

**Table 3.** Sensitivity, specificity and predictive values of morphologic assessment in discriminating oncocytic tumors into benign, borderline and malignant categories.

**A. Distribution of Actual and Predicted by pathology diagnoses**
|  |  | Actual (Cytogenetic) |  |  | TOTAL |
| --- | --- | --- | --- | --- | --- |
|  |  | Positive |  | Negative |  |
|  |  | ChRCC | RO-variant | RO |  |
| Predicted by pathology | ChRCC | 6 | 0 | 1 | 7 |
|  | HOCT | 3 | 5 | 3 | 11 |
|  | RO | 1 | 2 | 17 | 20 |
| TOTAL |  | 10 | 7 | 21 | 38 |

| Diagnostic test characteristics * |  | Actual (Cytogenetic) |  |  | TOTAL |
| --- | --- | --- | --- | --- | --- |
|  |  | Positive |  | Negative |  |
|  |  | ChRCC | RO-variant | RO |  |
| Predicted by pathology | ChRCC | TP (14) |  | FP (4) | 18 |
|  | HOCT |  |  |  |  |
|  | RO | FN (3) |  | TN (17) | 20 |
| TOTAL |  | 17 |  | 21 | 38 |

\* Sensitivity 82%, Specificity 81%, PPV 78%, NPV 85%, Accuracy 82%

| Diagnostic test characteristics * |  | Actual (Cytogenetic) |  |  |  |
| --- | --- | --- | --- | --- | --- |
|  |  | Positive | Negative |  |  |
|  |  | ChRCC | RO-variant | RO | TOTAL |
| Predicted by pathology | ChRCC | TP (6) | FP (1) |  | 7 |
|  | HOCT | FN (4) | TN (27) |  | 31 |
|  | RO |  |  |  |  |
| TOTAL |  | 10 | 28 |  | 38 |

\* Sensitivity 60%, Specificity 96%, PPV 86%, NPV 87%, Accuracy 87%

We performed detailed analysis of diagnostic test characteristics of immunomorphologic assessment by compiling 2 × 2 tables with actual (cytogenetic) and predicted (pathologic) results. If RO considered the only true benign cytogenetic diagnosis (ChRCC plus RO-variant lumped as non-benign), pathologic prediction had sensitivity of 82%, specificity of 81%, positive predictive value (PPV) 78%, negative predictive value (NPV) 85% and accuracy 85%. If ChRCC considered the only true malignant cytogenetic diagnosis (RO plus RO-variant lumped as benign), pathologic prediction showed much lower sensitivity of 60% combined with much higher specificity of 96% and improved PPV (86%), NPV (87%) and accuracy (87%). Using only two immunomorphologic diagnostic categories (RO and ChRCC) will result in either overcalling or undercalling actual malignant tumors in up to 40% cases. On the other hand, tumors with intermediate/borderline features could be reliably classified by cytogenetic profiling into ChRCC, RO or RO-variant subtypes.

## DISCUSSION

Distinguishing renal oncocytomas (RO) from eosinophilic ChRCC using only histology and immunostaining is challenging. Herein we studied 38 oncocytic tumors with features intermediate between classic RO and the eosinophilic variant of ChRCC. Genomic profiling allowed classification of all cases into RO, RO-variant or ChRCC categories. Cytogenetic RO consisted of two categories: 48% with normal numeric chromosomal status and 52% with loss of chromosome(s) 1p, X or Y. The cytogenetic RO-variant group had additional chromosomal losses [-9q, –14 (2 cases), –13] and chromosomal gains [+1q (2 cases), +4, +7 (2 cases), +13, +19, +20, and +22]. Cytogenetic ChRCC could be divided into two distinct subtypes: 40% had hypodiploidy due to numerous losses of portions or entire chromosomes 1, 2, 6, 10, 11, 14, 16, 17, and 60% had multiple relative chromosomal losses due to hypotetraploidy.

Cytogenetic RO, RO-variant and ChRCC cases had similar clinical features with no statistically significant differences in patient demographics, tumor size, stage or outcomes, although the number of events and length of follow-up were modest. Interestingly, we found significant differences in tumor architectures in the three cytogenetic subtypes. The architectural differences were in histological patterns (organoid/nested, tubulocystic and solid/confluent), the quality of stroma (fibromyxoid and edematous), nuclear wrinkling (in >5% of cells), perinuclear halos and well-defined cell borders with clear cytoplasm (p<0.05). The frequency of these features was more pronounced in cytogenetic RO’s and ChRCC’s, whereas RO-variant group positioned in between except for tubulocystic architecture which was least uncommon in RO-variant compared to ChRCC or RO cases. Features such as calcifications/osseous metaplasia, hemorrhage, cellular pleomorphism, bi/multinucleation, presence of mitoses, fat or vascular invasion were non-discriminatory (Table 2). A recent large survey also suggested that in oncocytic tumors certain morphologic features are more frequently associated with malignancy and should be used for triaging cases for cytogenetic testing to rule out ChRCC, especially in biopsies [4].

Immunostaining in our study showed statistically significant difference between study groups only for CK7 (p<0.05), similar to previous studies [4, 17, 19, 20]. S100A1 was diffusely expressed in the vast majority of RO and RO-variant cases, but didn’t reach statistical significance arguing against using this analyte for differential diagnoses with eosinophilic variant of ChRCC.

Morphologic features correlated with cytogenetic subtype in 85% of RO’s. One case (5%) had a genetic profile of ChRCC and two cases had profiles of RO-variant cytogenetics (10%). Morphologic prediction of ChRCC was accurate in 86% cases having ChRCC cytogenetic profile while one case had a genetic profile consistent with RO (14%). Similarly, in the TCGA study four of nineteen eosinophilic ChRCC cases (21%) had no copy number alterations and should be reclassified as RO [12]. However, our morphologic prediction of cytogenetic ChRCC or RO would miss up to 40% cases if we did not have a third group with mixed morphologic features (Table 3). This third group, designated as hybrid oncocytic/chromophobe tumor (HOCT), was cytogenetically very heterogeneous and classified into cytogenetic RO, RO-variant or ChRCC in 27%, 46% and 27% cases, respectively. Therefore, our study strongly argues in favor of using a borderline or intermediate diagnostic category similar to conclusion of two recent studies [4, 15].

In the past HOCT may have been diagnosed as oncocytoma based on similarities in morphology, ultrastructure and molecular features [21]. Although loss of chromosome 14 has alterations and should be reclassified as RO [12]. However, our morphologic prediction of cytogenetic ChRCC or RO would miss up to 40% cases if we did not have a third group with been reported in RO [8, 22], monosomy 14 was found more frequently with additional chromosomal losses and gains in RO-variant than in RO in this study (Figure 2). Some recent studies also showed HOCT is a genetically heterogeneous group of tumors for which genomic profiling can help in the classification. Fourteen sporadic HOCT cases analyzed by FISH showed recurrent monosomy 20 in 50% cases and random multiple chromosomal gains and losses of chromosomes 1, 2, 6, 9, 10, 13, 17, 21 and 22 [23], and should be reclassified as ChRCC. Twelve HOCT cases studied by French group using array CGH showed no CNA in 58% cases and a small number of random chromosomal losses and gains in remaining 42% cases involving 1p, 3q, 5p, 7p, 10 and 18 [24]. This study suggested that HOCTs are not true hybrid tumors and might be more appropriately classified as ROs. Recent molecular study of 27 HOCT by MD Anderson group [15] reported 40% of tumors without CNA which should be reclassified to RO and remaining 60% with either loss of chromosome 1 or sex chromosome, and segmental losses of 1p, 2q, 5p, 6q, 22q and polysomy 7. They concluded that HOCT occupies an intermediate cytogenetic position between RO and ChRCC, clustering closer to RO than to ChRCC based on RNA transcript data. Similarly, our study showed that RO-variant was enriched in the HOCT cohort (46%) and has genetic profiles similar to RO, which was characterized by typical RO chromosomal loss –1/1p-, –14, –X, and –Y with a few additional copy number aberrations and mostly gains such as gain of 1q and 7 (Figure 2). Our study showed that the genomic profile of RO-variant was quite different from ChRCC genetic profile of hypodiplody with chromosomal losses –1, –2, –6, –10, –13, –17, –21, –Y in combination [7]. To summarize, tumors with intermediate/borderline features (HOCT) could be reliably classified by cytogenetic profile into ChRCC, RO or RO-variant subtypes.

Although hybrid oncocytic/chromophobe tumors (HOCT) are commonly associated with Birt-Hogg-Dube syndrome [5, 25], whether sporadic HOCTs are a pathologically and genetically distinct entity remains a point of debate [3, 6, 9, 23, 26]. Some believe that HOCT may represent a morphologic variant of RO with excellent outcome [24, 27], an intermediate stage of stepwise progression from RO to ChRCC [21, 28]. Others regard HOCT as a unique entity with metastatic potential based on chromosomal and molecular alterations that are not seen in typical ROs or ChRCCs [15]. Interestingly, syndromic HOCTs are multifocal with mosaic patterns of RO and ChRCC-like zones, whereas in sporadic HOCTs mostly have an ambiguous intermediate morphology similar to our findings [15, 23–25, 29]. Future study with larger sample size and longer follow-up data will be helpful to sort out the association between the genetic profiles and patient outcome.

Hypodiploidy with multiple chromosomal losses of Y, 1, 2, 6, 10, 13, 17 and 21 is a hallmark of ChRCC [7], however, some studies reported ChRCC cases with gains of chromosomes 4, 7, 12, 14, 15, 18q, 19 and 20 at lower frequencies of ChRCC cases [12, 30–34]. These cases were interpreted as hyperdiploid with multiple chromosomal gains, but they most likely resemble hypotetraploid ChRCC (4n) described in this study because the chromosomes with three or four copies in the six hypotetraploid ChRCC cases of this study were also most frequently chromosomes 4, 7, 12, 14, 15, 16, 18, and 20 (Supplemental Table 1, Figure 2). Given the SNP pattern and copy number status for each chromosome examined as described in the results section, these ChRCC cases had in fact doubled hypodiploid genomes with numerous chromosomal losses relative to tumor polyploidy status (4n) including most frequently two-copy loss of chromosomes Y, 1, 2, 6, 10, 13, and 17 (Table 1, supplemental Table 1, Figure 2). Our intriguing observation of doubled hypodiploidy (4n) with relative chromosomal losses explained the discrepancy found in the previous studies that some ChRCC had gains of chromosomes 4, 7, 12, 14, 15, 18q, 19 and 20 instead of multiple chromosomal losses. While majority of ChRCC had hypodiploid genomes, some had doubled hypodiploid genomes (4n, hypodetraploidy), and both had same set of multiple chromosomal losses of Y, 1, 2, 6, 10, 13, 17 and 21. Doubled hypodiploidy was also supported by several earlier studies. A study using flow cytometry and quantitative image cell analyses of a series of ChRCC showed portion of doubled hypodiploid nuclei in ChRCC with their combined DNA content essentially similar to that of single hyperdiploid nuclei, suggesting polyploidy resulting from the fusion/doubling of these nuclei [35]. Several cytogenetic studies revealed nine ChRCC cases having hypotetraploid karyotypes with multiple chromosome losses, and four of nine cases also had a hypodiploid stem line [19, 36, 37]. More recent study of ChRCC showed imbalanced chromosome duplication (ICD, duplication of ≥3 chromosomes) in 25% of metastatic ChRCC cases [32] and in one case of HOCT with liver metastasis [15]. These findings demonstrated clonal evolution and polyploidization, and were associated with more aggressive behavior. This phenomenon may indeed be the basis for tumor cell heterogeneity in ChRCC with eosinophilic features, such as separate coexisting clones within the same tumor, and polyploidization as a compensatory mechanism to maintain the genetic balance in near haploid/hypodiploid cells. Our study also shows that hypotetraploid (doubled hypodiploidy) with multiple relative losses of this same set of chromosomes is a common phenomenon that is enriched in eosinophilic ChRCC (60%).

In summary, genomic profiling should be used to reliably categorize oncocytic tumors with ambiguous morphology and immunoprofiles into RO, RO-variant and ChRCC. Specific architectural features and status of CK7 expression correlate with cytogenetic-based categories-RO, RO-variant and ChRCC tumors. The histological entity HOCT is a heterogeneous group enriched for cytogenetic RO-variant and equal chance of cytogenetic RO or ChRCC.

We also found that chromosome ploidy status has a strong correlation with histologic subtype. Double hypodiploidy (by whole genome endoduplication) is a common phenomenon in eosinophilic ChRCC.

## Data Availability

Data referred in the manuscript is available upon request

---

**Supplemental Table 1.** Detailed cytogenetic profiles of thirty-eight oncocytic tumors.

| Case ID<br>(numbers) | PathDX | GeneticDx | Ploidy<br>(F/M) | 1 copy<br>(loss) | 2 copies with LOH<br>(loss in 4n) | 2 copies without LOH<br>(loss in 4n) | 3 copies<br>(gain in 2n, loss in 4n) | 4 copies<br>(no loss in 4n) |
| --- | --- | --- | --- | --- | --- | --- | --- | --- |
| 16-20,25,32 (7) | RO | RO | 2n |  |  | normal |  |  |
| 8,21,29 (3) | HOCT | RO | 2n |  |  | normal |  |  |
| 14,26 (2) | RO | RO | 2n | 1 |  |  |  |  |
| 12,13,15,30 (4) | RO | RO | 2n | 1,Y |  |  |  |  |
| 33 (1) | RO | RO | 2n | 1,X |  |  |  |  |
| 31 (1) | RO | RO | 2n | 1p<sup>p</sup> |  |  |  |  |
| 6 (1) | RO | RO | 2n | 1p<sup>p</sup>,Y |  |  |  |  |
| 24 (1) | RO | RO | 2n | 1p<sup>p</sup> |  |  |  |  |
| 7 (1) | ChRCC | RO | 2n | 1p<sup>p</sup> |  |  |  |  |
| 11 (1) | RO | RO-variant | 2n | 9q<sup>p</sup> |  |  |  |  |
| 28 (1) | RO | RO-variant | 2n | 1,Y |  |  | 19, 22 |  |
| 22 (1) | HOCT | RO-variant | 2n | 1p |  |  | 1q |  |
| 27 (1) | HOCT | RO-variant | 2n | 1p<sup>p</sup>,1q<sup>p</sup> |  |  | 1q<sup>p</sup> |  |
| 4 (1) | HOCT | RO-variant | 2n | 1,14,X |  |  | 20 |  |
| 5 (1) | HOCT | RO-variant | 2n | 13,14,Y |  |  | 4,7 |  |
| 23 (1) | HOCT | RO-variant | 2n |  |  |  | 7,13 |  |
| 9 (1) | HOCT | ChRCC | 2n | 1,2q<sup>p</sup>,6q |  |  |  |  |
| 35 (1) | HOCT | ChRCC | 2n | 1,2q,7,10,11q<sup>p</sup>,16 |  |  |  |  |
| 10 (1) | ChRCC | ChRCC | 2n | 1,2,5,6,9q<sup>p</sup>,10,13,17,X |  |  |  |  |
| 34 (1) | RO | ChRCC | 2n | 1p,2p<sup>p</sup>,6q<sup>p</sup>,8p<sup>p</sup>,9p<sup>p</sup>,11q<sup>p</sup>,14q<sup>p</sup> |  |  | 3q<sup>p</sup> |  |
| 1 (1) | HOCT | ChRCC | 4n (F) | X | 1,6,8,9,11,14,15,18,22 | 2,19 | 3,4,10,16,17,21 | 5,7,12,13,20 |
| 2 (1) | ChRCC | ChRCC | 4n (F) |  | 1,11,13,20 | 2,5,8,9,10,17,19 | 3,6,12,14,16,18,21,X | 4,7,15,22 |
| 3 (1) | ChRCC | ChRCC | 4n (M) | 8p,11q<sup>p</sup>,14,18q<sup>p</sup>,X<sup>p</sup>,Y | 1p,2q<sup>p</sup>,3,4,6,10,13 | 2p<sup>p</sup>,8q,9,11p<sup>p</sup>,15,17,18p<sup>p</sup>,19,21,22 | 1q,5 | 7,12,16,20 |
| 36 (1) | ChRCC | ChRCC | 4n (F) |  | 2,8,13,17 | 1,6,10 | 3,11,19,21,X | 4,5,7,9,12,14,15,16,18,20,22 |
| 37 (1) | ChRCC | ChRCC | 4n (M) | Y | X | 1,2,3,5,6,9,10,17,19,20,21 | 8,11,12,13,14,15,16,18 | 4,7,22 |
| 38 (1) | ChRCC | ChRCC | 4n (M) | Y | 2,6,13,17,X | 1,5,10 | 3,12,14,19,21 | 4,7,8,9,11,15,16,18,20,22 |

RO – oncocytoma, ChRCC – chromophobe renal cell carcinoma, HOCT – “hybrid” oncocytoma/chromophobe tumor; 2n – diploid, 4n – tetraploid, p – short arm of chromosome, q – long arm of chromosome, <sup>p</sup> – portion of the chromosomal arm (see supplemental table 1 for details).

